# Prevalence and Associated Risk Factors for Early Childhood Caries in Central Africa Sub-Region: A Systematic Review and Meta-analysis

**DOI:** 10.64898/2026.07.10.26357776

**Authors:** Oluwakemi Ojulowo, Chidera Okoli, Folahanmi Akinsolu, Adebola Ehizele, George Uchenna Eleje, Qudus Olajide Lawal, Richard Oveh, Agbor Micheal Ashu, Oliver C. Ezechi, Moréniké Oluwátóyìn Foláyan

**Affiliations:** Department of Paediatrics, Federal Teaching Hospital, Ido-Ekiti, Ekiti State, Nigeria; Department of Biochemistry, Chukwuemeka Odumegwu University, Uli, Anambra state, Nigeria; Department of Public Health, Faculty of Basic Medical and Health Sciences, Lead City University, Ibadan 110115, Nigeria; Department of Periodontics, School of Dentistry, College of Medical Sciences, University of Benin, Benin City, Nigeria; Department of Obstetrics and Gynaecology, Nnamdi Azikiwe University, Awka, Nigeria; Irrua Specialist Teaching Hospital, Irrua, Nigeria; University of Delta, Agbor, Delta State, Nigeria; Faculty of Dentistry, Université des Montagnes, Cameroon; Department of Child Dental Health, Faculty of Dentistry, College of Health Sciences, Obafemi Awolowo University, Ile-Ife, Nigeria

**Keywords:** Central Africa, Early Childhood Caries, ECC prevalence, Risk factors

## Abstract

**Background:** Children in Central Africa face compounded vulnerabilities, yet no comprehensive review has synthesized evidence on ECC prevalence or risk factors. This study aims to determine the prevalence and identify risk factors associated with early childhood caries (ECC) in the Central Africa Sub-region.

**Methods:** This systematic review and meta-analysis was registered with the International Prospective Register of Systematic Reviews (PROSPERO) under the ID CRD420251019424. A comprehensive literature search was conducted across PubMed, African Journals Online, Scopus, Web of Science, CINAHL, and ProQuest as well as through Google Scholar and other grey literature sources, with no language restrictions. Eligible studies were limited to cross-sectional, cohort, case-control, and clinical trials that reported baseline data on the prevalence and risk factors of ECC in the Central Africa sub-region among children under the age of six. There was no restriction on publication date. Exclusion criteria included letters to the editor, commentaries, studies without accessible full texts, systematic reviews, reviews lacking original data, studies with overlapping data already included, and non-peer-reviewed sources such as books. Study quality was assessed using the Joanna Briggs Institute Critical Appraisal Tool. A random-effects meta-analysis was conducted using Review Manager 5.4.1, and heterogeneity was assessed using the I² statistic.

**Results:** Five studies met the eligibility criteria, encompassing 640 participants. Three of the studies were from Cameroon, and one from the Democratic Republic of Congo and the Central African Republic, respectively. One study that did not report the prevalence of ECC and another non-peer-reviewed publication were excluded from the meta-analysis. The prevalence of ECC in the region ranged between 29.4% and 80% with a pooled prevalence of 57% (95% CI: 22–92%; I² = 98%). Identified risk factors for ECC were related to oral hygiene behaviours, diet and feeding practices, parental/caregiver factors, and access to dental care.

**Conclusions:** ECC is highly prevalent in Central Africa, driven by preventable factors. Urgent region-specific policies and programs are needed to improve preventive services and address data gaps across this under-resourced subregion.

Clinical trial registration number: Not applicable

## Introduction

Early Childhood Caries (ECC) is defined as the presence of one or more decayed (either non-cavitated or cavitated), missing (due to caries), or filled surfaces in any primary tooth of a child younger than six years old. [1] ECC represents a significant global public health problem. It can severely impact a child’s quality of life, affecting their ability to chew food, which contributes to malnutrition, recurrent pain, oral infections, poor sleep, low self-esteem, and impaired social interactions. [1–6]

Globally, ECC affects approximately 49% of young children, compared with 42% in Africa overall. [7] However, the rising burden of dental caries in African countries remains a major concern. [8] This increase occurs against a backdrop of longstanding challenges, including limited access to oral healthcare services [9], inadequate public health policies [10, 11], poor oral health literacy [12, 13], and a high consumption of sugary diets associated with urbanization and dietary transitions. [14, 15] Socioeconomic disparities [16, 17], shortages of trained dental professionals and a lack of school-based preventive programs [18] further compound these issues. Together, these factors demand evidence-based, strategic interventions to curb the growing burden of ECC.

Central Africa requires particular attention due to the region’s consistently poor health profile. [19] Countries in this sub-region have some of the lowest national incomes, with an average GDP per capita of approximately $1,500, which is significantly below the continental average of $2,500. [20] The Gini index often exceeds 40, indicating significant economic inequality and barriers to healthcare access. [21] Life expectancy also remains low, averaging 55 years compared to the African average of 64 years, with some countries, such as the Central African Republic, reporting life expectancy as low as 54 years. [22]. These economic and health disparities likely contribute to an elevated risk of ECC. [7]

Despite this, no comprehensive review has examined the prevalence and risk factors for ECC in Central Africa. This lack of evidence makes planning and resource allocation for prevention and control programs difficult. To address this gap, this study systematically reviews existing research to estimate the prevalence of ECC and identify associated risk factors in the Central African sub-region.

## Methods

### Protocol and Registration

The protocol for this systematic review and meta-analysis was registered with the International Prospective Register of Systematic Reviews (PROSPERO) under the number CRD420251019424, published publicly on April 4, 2025. The study was reported following the Preferred Reporting Items for Systematic Reviews and Meta-analyses (PRISMA) statement and checklist [23].

### Research Questions

This systematic review and meta-analysis aimed to answer the following research questions: (i) What is the prevalence of ECC in the Central Africa sub-regionand (ii) What risk factors are associated with ECC in the Sub-region?

### Data Sources and Search Strategy

The initial search was conducted in April 2025 using six electronic databases (PubMed, African Journal Online, Scopus, ProQuest, Web of Science, and Cinahl), as well as grey literature sources. The search was conducted using the following key terms: “prevalence,” “epidemiology”, “incidence”, “early childhood”, “preschool” “infant”, “dental caries,” “caries,” “tooth decay,” early childhood caries”,” “preschool,” “early childhood,” “ Search terms were tailored to the specific requirements of each database. Boolean operators (AND, OR) and truncation were applied to refine the search results. The search string for each database is in Supplemental File 1.

In addition, to enhance the breadth of sources retrieved, the phrase “caries and prevalence and children and the name of each country in the Central Africa sub-region” was used as a search term in Google Scholar. Google Scholar, an internet-based search engine, was utilized to enhance coverage and ensure the inclusion of relevant research outputs from the region. [24].

The reference lists of the included studies were manually reviewed to identify any additional relevant papers. Dental schools and oral health institutions listed with the World Health Organization African Region were contacted for possible grey materials. An internet search was also conducted to locate grey literature, such as conference proceedings and reports, to capture pertinent materials not indexed in conventional academic databases. The search was conducted without any language restrictions to maximize inclusivity.

### Eligibility Criteria

This review included only primary studies (cross-sectional, cohort, case-control, and clinical trials) that provided relevant baseline data and reported on the prevalence and risk factors associated with ECC in countries in the Central Africa Sub-region. Studies conducted in Cameroon, the Central African Republic, Chad, Congo, the Democratic Republic of the Congo, Equatorial Guinea, Gabon, and São Tomé and Príncipe, which included children under 6 years old, were eligible for inclusion in the study. There was no restriction on the date and language of the study. Letters to the editor, commentaries on studies, primary studies whose full articles cannot be accessed, systematic reviews, reviews devoid of primary data, studies with overlapping data from other included studies, and other data sources, such as books, were excluded.

### Selection of Data Sources

Eligibility screening of studies was guided by the CoCoPop framework—Context, condition, and Populations. [25] . The population of interest was children aged 0-71 months living in any of the countries in Central Africa, regardless of sex. The condition examined was dental caries. The PEO framework was used to assess for risk factors of ECC within this group.

Articles obtained from the databases were downloaded into the Zotero reference manager and imported into Rayyan. Duplicate articles were removed using the “find duplicate” function and manually when encountered during the review. Two independent reviewers (OBO and CGO) screened the titles and abstracts based on the eligibility criteria. The studies included were based on an agreement between the two reviewers. Where conflict existed, a third reviewer (MA) was consulted, who screened the articles and made the decision. Full-text articles of all studies that were included following the two-stage screening process were obtained and reviewed by two independent reviewers (OBO and CGO). The inter-rater reliability score was determined using the Kappa statistic.

### Data Collection Process

Data extraction focused on key information related to the study participants, the exposure of interest, and the outcomes relevant to the review objectives. Two reviewers (OBO and CGO) independently carried out the data extraction process using a pilot-tested extraction form to ensure consistency and reliability. This method reflects established best practices in systematic reviews, as independent extraction by multiple reviewers reduces the risk of errors and bias [26, 27].

### Data Extraction Process

A spreadsheet was created on Microsoft Excel to extract relevant variables from the articles. Details such as the authors’ names and publication years were documented. In addition, relevant information on each study’s design (cross-sectional, ecological, case control, and cohort), sample size, study location (country, rural or urban area), and participant characteristics (sample size, age range, sex distribution, and other distinguishing features). Data about the prevalence of ECC and identified ECC risk factors, including possible preventive behaviour (oral hygiene status, frequency of tooth brushing, use of fluoridated toothpaste, daily intake of refined carbohydrates, and dental service utilization), were extracted. Disagreements that arose during the data extraction were again reviewed and resolved by the senior author (MOF).

### Risk of Bias Assessment

Each of the studies that met the eligibility criteria was assessed for the risk of bias using the Joanna Briggs Institute critical appraisal tool for risk of bias assessment. [28]. Each of the eight items on the JBI checklist was evaluated using four response options: ‘Yes’, No’, ‘Unclear’, and ‘Not applicable’. A score of 2 points was assigned for ‘Yes’, 1 point for ‘No’, and 0 points for both ‘Unclear’ and ‘Not applicable’. The total quality score for each study ranged from 0 to a maximum of 16. Studies scoring between 12 and 16 were classified as ‘high quality’, those scoring 9 to 11 as ‘moderate quality’, and those with a score of 8 or below as ‘low quality’. The results were presented in a tabular format to enable comparison across studies and ensure that the synthesis was informed by relevant and high-quality evidence. Disagreements that arose during bias assessments were again reviewed and resolved by the senior author (MOF).

### Assessment of Publication Bias

Symmetry of the funnel plot was used to assess the publication bias of the articles included in the systematic review. Generally, publication bias was not assessed when the included studies had fewer than 10 participants.

### Sensitivity Analysis

A sensitivity analysis was conducted to assess the impact of data changes on the overall results and to identify potential sources of heterogeneity. This involves leaving out one sensitivity analysis. The goal was to determine whether the main findings are robust to these changes, thereby ensuring the reliability of the conclusions.

### Data Analysis

Studies with moderate and low risk of bias were included in the meta-analysis for the synthesis of results. Data were analyzed using RevMan 5.4.1 (The Nordic Cochrane Center, Copenhagen, Denmark). To estimate the prevalence of ECC, data were pooled using a 95% confidence interval (CI) as the measure of effect size. The Generic Inverse Variance method was employed to calculate the combined effect. In this approach, the rate difference (RD) and its standard error correspond to the effect of a single rate and its standard error. Heterogeneity among studies was evaluated using Cochran’s Q test and Higgins and Thompson’s I² statistic. A p-value less than 0.05 indicated statistical significance for the Q test, while an I² value greater than 50% denoted significant heterogeneity. For all other tests, except heterogeneity testing, p-values less than 0.05 were considered statistically significant.

## Results

A total of 74 articles were retrieved from searches conducted. All records were imported into Zotero and then uploaded to Rayyan for screening. Nine duplicates were removed before the screening process. During title and abstract screening, 58 articles were excluded because they were not conducted in countries in Central Africa or they did not have data on ECC. Six articles met the eligibility criteria. However, one article could not be retrieved. Of the five eligible studies, one was excluded from the meta-analysis because it did not have data on the prevalence of ECC, and another was removed because it was not peer reviewed. The interrater reliability was high, with a Cohen’s kappa of 0.75 [29]. Figure 1 presents the PRISMA flow diagram of the screening process, and Table 1 summarizes the characteristics and key findings of the included studies.

**Figure 1:**
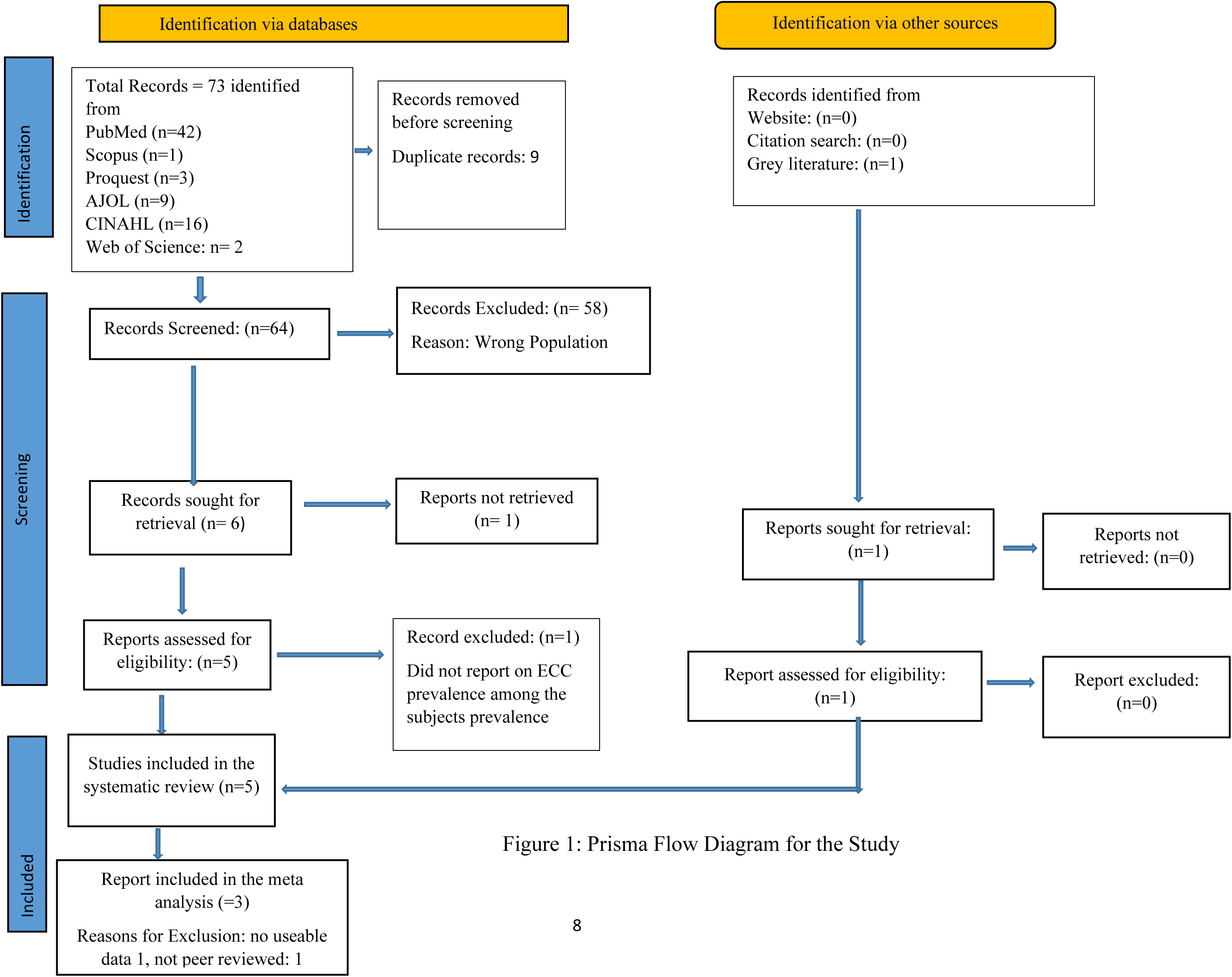
Prisma Flow Diagram for the Study

**Table 1.** Characteristics and Some Findings of the Included Studies.

| Name of Author, Year of Publication | Country of study | Study location | Study design | Sample size | Age range and sample size by sex | Dental caries diagnostic criteria | Dental caries risk indicators | Study Quality | Prevalence of dental caries | Study findings |
| --- | --- | --- | --- | --- | --- | --- | --- | --- | --- | --- |
| Lamloum et al, 2025 [34] | Central Africa Republic | School | Cross sectional | 288 | 5–6-year-old<br>Male:153<br>Female:135 | dmft | Sex, frequency of brushing, frequency of snack intake, dental visits, number of meals per day, and use of toothbrush | High | 48% | The mean dmft was $2.19 \pm 2.95$. Caries was significantly associated with peri-urban school location ($p = 0.01$), but not with sex, parental education, or eating habits. Over half of the participants had never accessed dental care. |
| Aimond et al, 2023 [30] | Cameroon | Community | Cross sectional | 45 | 3–18-year-old<br>Male: 27<br>Female:18 | ICDAS | Frequency of tooth brushing, frequency of drinking soda, frequency of eating candy, and dental visits | Moderate | 62.2% | 98% consumed candies, and this increased with age. Gingivitis affected 27.2%, moderate tartar was found in 41%, and 28% had poor oral hygiene. 95.1% had never seen a dentist. |
| Keunang, 2023 [31] | Cameroon | Community | Cross sectional | 43 | 3–6-year-old<br>Male:20<br>Female: 23 | dmft | Frequency of tooth brushing, consumption of sweetened drinks or food before sleep, bottle or breast feeding before sleep, and dental visits | Moderate | 41.0% | The mean dmft was 1.25. More females had dental caries than males. Poor oral hygiene was a risk factor for dental caries, and about 81% of the children had never visited the dentist. |
| Songo et al, 2013 [33] | Democratic Republic of Congo | Hospital | Cross sectional | 158 | 4–6-year-old<br>Male:79<br>Female: 79 | dmft | Sex, level of education of both parents, dental history of parents, bottle feeding, frequency of sugar-containing meals and drinks | High | 80.0% | Girls had a lower risk of dental caries (OR 0.43). Higher risk was associated with eating more than two meals daily (OR 2.7), consuming multiple sugary drinks per day (OR 2.47), and eating sugary foods before bedtime (OR 3.39). Children of mothers with a history of dental caries had a reduced risk (OR 0.38). |
| Poni et al, 2023 [32] | Cameroon | Community | Cross sectional | 106 | 5-6 year old | dmft | Diet, frequency of tooth brushing, techniques used for tooth brushing, tooth brushing instruments, and aids. | Moderate | 29.24% | The mean dft index was 0.73. Only 2% consumed cariogenic foods. Toothbrushing was reported by 31% of children. No association was found between diet and dental caries. Only 13% had ever visited a dentist. |

### Characteristics of Included Studies

Of the five studies included in the systematic review, three were from Cameroon. [30–32], one from the Democratic Republic of the Congo [33], and one from the Central African Republic [34]. No studies were retrieved for Chad, Equatorial Guinea, Congo, Gabon, and São Tomé and Príncipe.

The studies were published between 2013 and 2025, with four studies published between 2023 and 2025 [30–32, 34]. All the studies were cross-sectional in design. Three studies were community-based [30–32], one school-based [34], and one was hospital-based [33]. The sample size for study participants ranged from 43 [31] to 556 [33], with a total sample of 1,165 participants. Of these, 640 were children under six years old. Four studies [30, 31, 33, 34] gave the distribution of children under six years by sex (279 males and 255 females), and one study did not report the sex distribution of participants less than 6 years old [32].

Four studies utilized the ‘dmft’ index as a diagnostic tool [31–34] and one utilized the International Caries Detection and Assessment System (ICDAS) Grade [30]. Only one study exclusively examined children under six years old [31]. The other studies included preschool children as part of studies in children aged three to eighteen years [30, 32–34].

### Study Findings on ECC in Central Africa

The reported mean ‘dmft’ scores ranged from 1.25 [30] to 2.76 [34], indicating moderate levels of untreated dental decay in the sampled populations. One study [32] reported an index of 0.735 among study participants, indicating a very low level of dental caries in that population.

### Risk Indicators for Early Childhood Caries

Table 2 highlights the risk factors analysed for ECC in the studies, and they include tooth brushing frequency [32, 34], intake of sugary foods and drinks, particularly candies [30, 32, 34], sweet snacks [34], and sodas [30], bedtime feeding with bottles [31] or breastfeeding [33], and fresh fruit consumption [33] that are considered individual-level factors. Family and household-level factors, including parental education [33], parental dental history [33], and bottle feeding [33]. At the structural and community level, access to dental care [32, 34], residential location [35], use of toothbrushes and fluoridated toothpaste [32, 33], nutritional patterns, including the type of diet [32], frequency of meal intake per day [34] and healthy meals [33].

**Table 2.** Emerging Thematic Clusters on Risk Indicators for ECC Studied in Central Africa.

| Thematic Cluster | Associated Variables |
| --- | --- |
| **Oral hygiene behaviours** | Frequency of brushing [30–32, 34], toothbrush use [32, 34], toothpaste use [32, 34] |
| **Diet and feeding practices** | Snack intake [33, 34], sugary drinks [31, 33], bottle/breastfeeding at night [31], candy [30], meal frequency [34] |
| **Parental/caregiver factors** | Education level [33], dental history [33] |
| **Access and environment** | Dental visits [30, 31, 34], housing location [30] |

#### Behavioural and dietary risk factors

Several behavioural factors were associated with increased risk for ECC. These include frequent snacking [34], consumption of sugar-containing drinks [33], candy consumption, and evening meals high in sugar [33]. Additionally, skipping breakfast was associated with an increased risk of caries, while regular breakfast intake had a protective effect [34]. Poor oral hygiene (lack of tooth brushing or use of fingers) was also a risk factor [31]. Reduced consumption of cariogenic foods like sweets and candies was noted among the population with a low ECC prevalence [32].

#### Socioeconomic and structural factors

Limited access to dental services was a consistent issue, with over 80%–95% of children having never visited a dentist [30–32]. A low prevalence of ECC was reported among the Baka Pygmies, who are nomadic, reside in camps with little access to cariogenic food [32].

#### Parental influence

Children whose mothers had experienced caries had lower odds of developing ECC themselves [33].

#### Sex differences

The findings on sex-related issues were mixed. Songo et al. [32] found that girls were at a lower risk of ECC, while Lamloum et al. [34] and Keunang [31] reported that females had a higher risk.

#### Risk of Bias Assessment

Two independent reviewers assessed the risk for bias for each study. Table 3 highlights the results of the risk of bias assessment. The eligible studies had low [33, 34] to moderate [30–32] risk of bias. The interrater reliability was 70% Supplementary Table 3 shows the interrater scores for the risk of bias assessment. When conflicts arose, they were resolved through discussion between the reviewers.

**Table 3.**
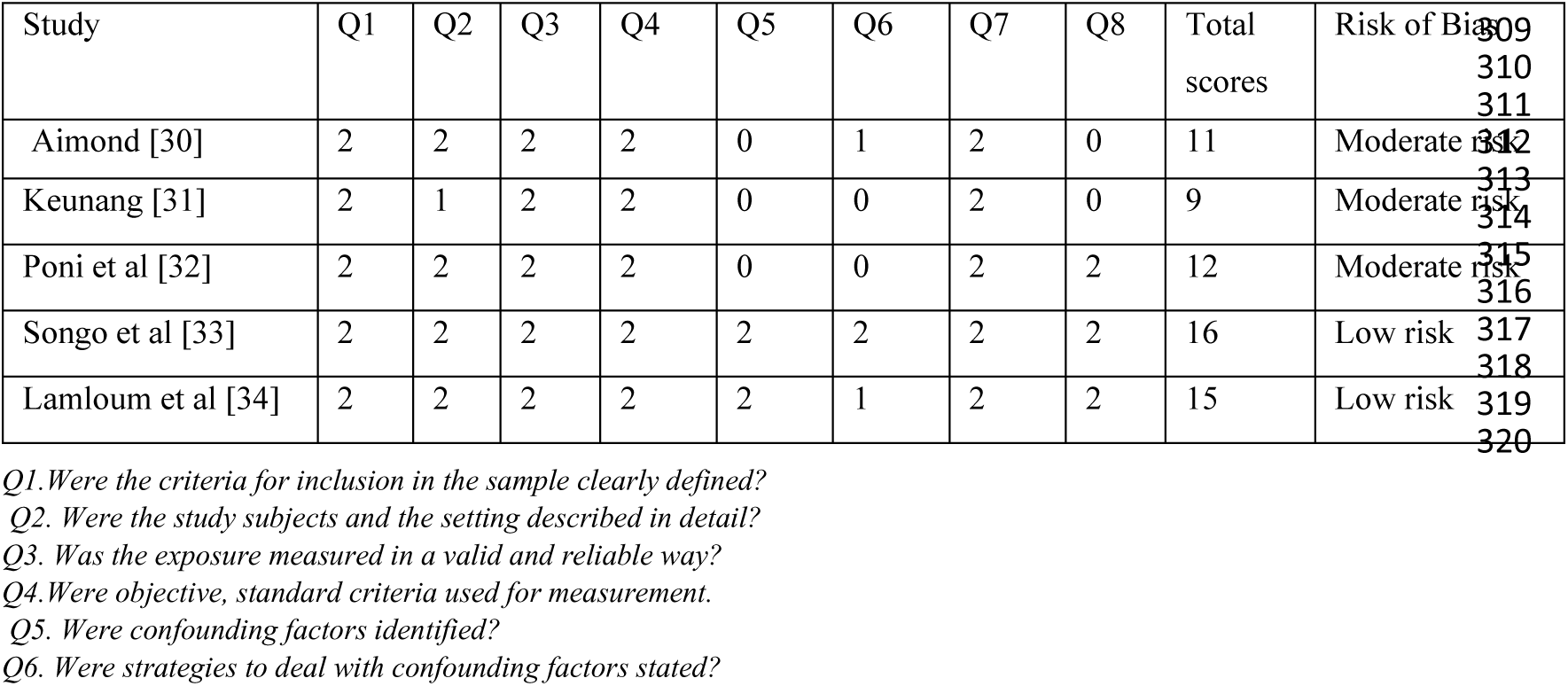
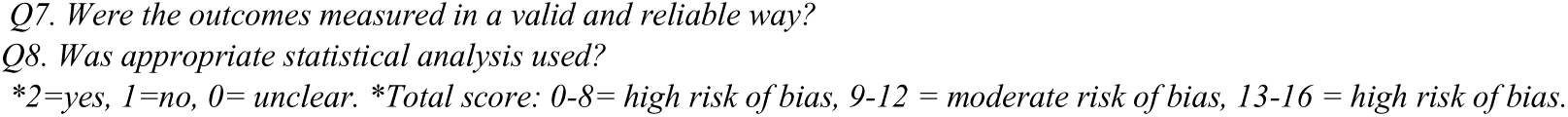
Risk of Bias Assessment for Each Study.

| Study | Q1 | Q2 | Q3 | Q4 | Q5 | Q6 | Q7 | Q8 | Total scores | Risk of Bias |
| --- | --- | --- | --- | --- | --- | --- | --- | --- | --- | --- |
| Aimond [30] | 2 | 2 | 2 | 2 | 0 | 1 | 2 | 0 | 11 | Moderate risk |
| Keunang [31] | 2 | 1 | 2 | 2 | 0 | 0 | 2 | 0 | 9 | Moderate risk |
| Poni et al [32] | 2 | 2 | 2 | 2 | 0 | 0 | 2 | 2 | 12 | Moderate risk |
| Songo et al [33] | 2 | 2 | 2 | 2 | 2 | 2 | 2 | 2 | 16 | Low risk |
| Lamloum et al [34] | 2 | 2 | 2 | 2 | 2 | 1 | 2 | 2 | 15 | Low risk |

Q1. Were the criteria for inclusion in the sample clearly defined?
Q2. Were the study subjects and the setting described in detail?
Q3. Was the exposure measured in a valid and reliable way?
Q4. Were objective, standard criteria used for measurement.
Q5. Were confounding factors identified?
Q6. Were strategies to deal with confounding factors stated?
Q7. Were the outcomes measured in a valid and reliable way? Q8. Was appropriate statistical analysis used? \*2=yes, 1=no, 0= unclear. \*Total score: 0-8= high risk of bias, 9-12 = moderate risk of bias, 13-16 = high risk of bias.

#### Pooled Prevalence of Early Childhood Caries in Central Africa

The prevalence of ECC reported by four of the five studies ranged from 29.4% [32] to 80% [33]. The prevalence of ECC was low among Baka pygmies, a nomadic community with rural life style. Figure 2 shows that the pooled prevalence of ECC was 57.0% (95% CI: 22%, 92%). In addition, the heterogeneity index of I^2^ = 98% (p=0.0001) indicated substantial heterogeneity among the included studies.

**Figure 2:**
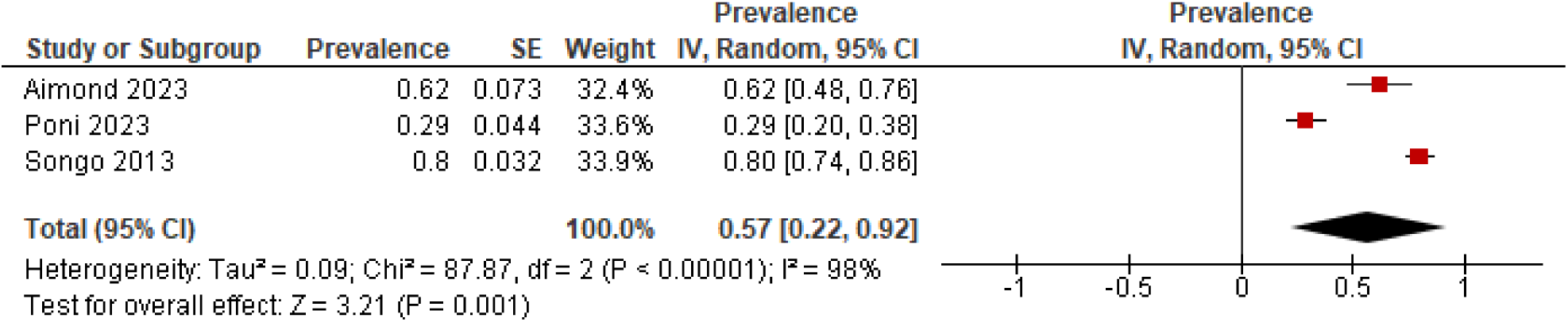
Pooled prevalence of ECC in Central Africa

**Figure 3:**
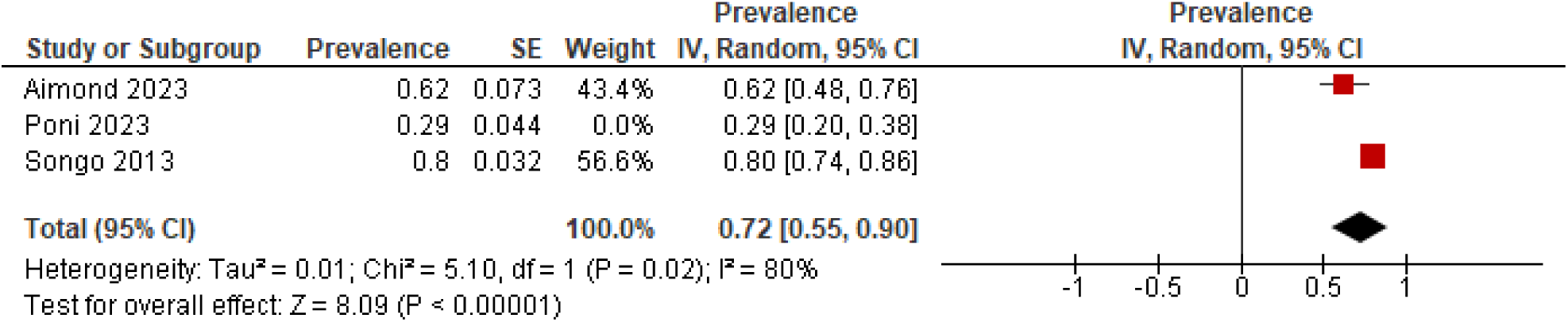
Leave-one-out sensitivity analysis of included studies

**Figure 4:**
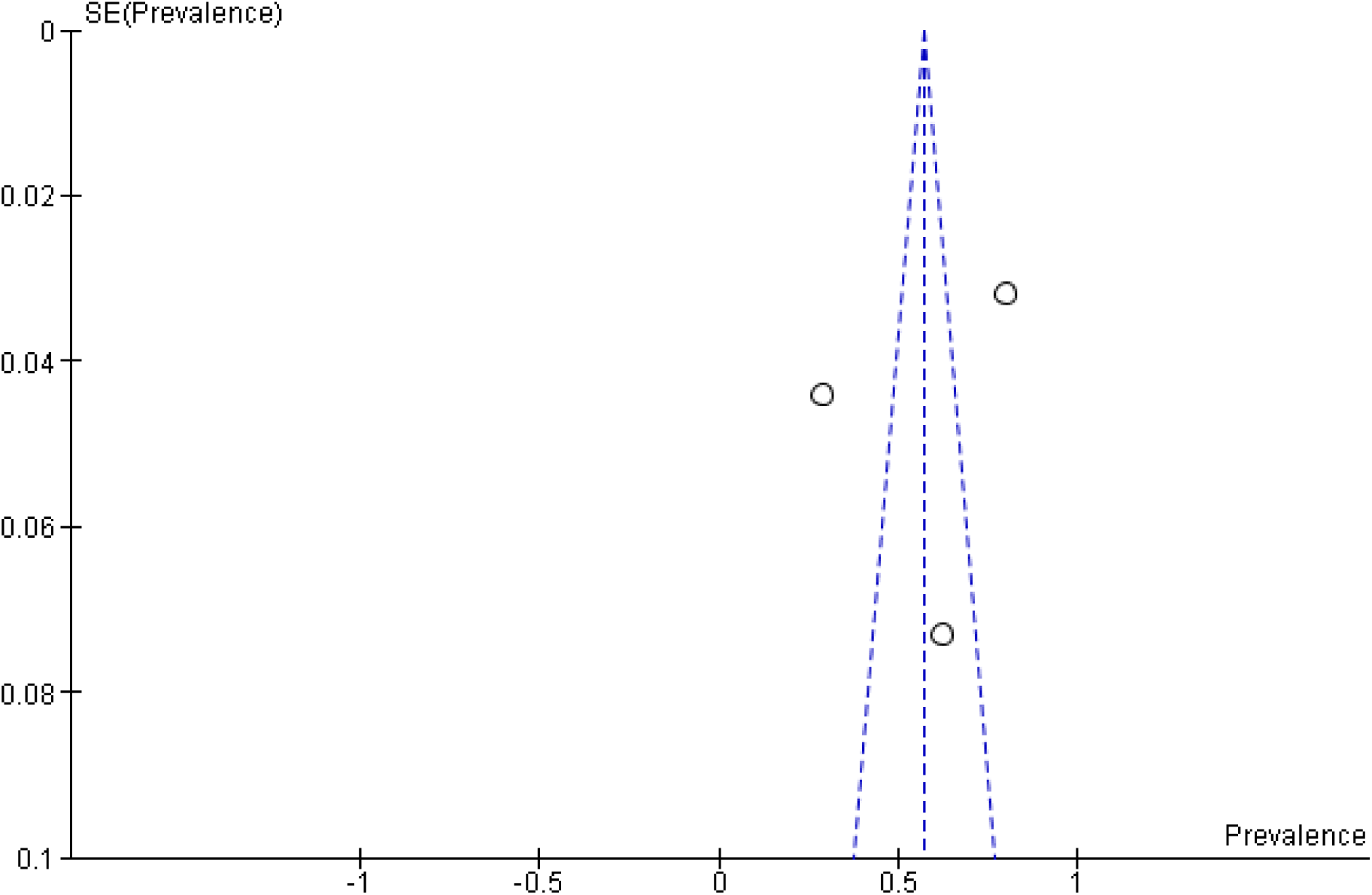
Funnel plot of included studies

#### Sensitivity Analysis

The leave–one–out sensitivity analysis showed that the pooled prevalence of ECC in Central Africa ranged from 0.45 to 0.72. Excluding Poni et al [32] increased the pooled prevalence (0.72 [0.55–0.90]) and reduced Tau² from 0.09 to 0.013 (I² 80 %), indicating this study as the main source of heterogeneity since it was conducted among a nomadic community. Exclusion of either Aimond et al [30] or Songo [33] had only modest effects on the pooled point estimate but left heterogeneity very high (I² > 93 %).

#### Publication Bias

A funnel plot of the studies included in the meta-analysis revealed an asymmetrical distribution of the studies across the funnel plot. This asymmetry is probably due to the low prevalence of ECC reported by Poni et al [32] and the limited number of studies included in the analysis not publication bias.

#### Risk Factors Associated with Early Childhood Caries

Two [33, 34] of the five studies included in this review, conducted bivariate analysis to determine risk factors for ECC among study participants with several risk factors analysed across the studies; however, the majority of the definitions for each of the factors analysed were not comparable across the studies. A meta-analysis of associated risk factors was not conducted due to the limited number of available studies.

## Discussion

This systematic review and meta-analysis provides the first comprehensive estimate of the prevalence of ECC in the Central Africa sub-region, showing that approximately 1 in 2 children under 72 months are affected. In addition, modifiable individual (behavioural and dietary risk factors), household (parental influence), and macro-level (socioeconomic and structural) factors were identified as risk factors for ECC.

A major strength of this review is its methodological rigour. The study followed PRISMA guidelines, employed a comprehensive search strategy that included grey literature, conducted independent dual screening, and assessed the risk of bias using the Joanna Briggs Institute tool. Additionally, it also addresses a significant data gap in a region where oral health research is limited.

Despite these strengths, several limitations should be noted. Only five studies met the inclusion criteria, with a sample size that limits the generalizability of the findings. Also, there was considerable heterogeneity among the included studies, though sensitivity analysis helped identify its source. Furthermore, only one study focused exclusively on <6-year-olds. In addition, five of the eight countries in the region (Chad, Congo, Gabon, Equatorial Guinea, and São Tomé and Príncipe), had no eligible data, suggesting gaps in surveillance and reporting that further limit regional representativeness and the generalizability of the study findings. Inspite of the limitations, the study highlights some critical findings.

First, compared to other regions globally, the burden of ECC in Central Africa is high. The pooled prevalence exceeds the WHO African regional average of 42% [7], and is higher than rates reported in Central and South America (34%), Europe (36%), and Asia-Oceania (52%) [7]. However, North America (57%) has comparable data, while the Middle East had a higher pooled prevalence of 72% [7]. This reflects a serious public health challenge for young children in the region, particularly considering the poor utilisation of dental services reported by most of the studies.

These findings also illustrate how biological risk factors intersect with broader social vulnerabilities. Consistent with the ecological model of oral health, individual behaviours, family circumstances, and structural determinants all shape the risk of ECC [18, 36]. Poor oral hygiene practices, frequent consumption of sugary foods and drinks, low use of dental services, and socioeconomic inequalities were the main ecological contributors identified in the included studies. Sex differences emerged, though findings were inconsistent. Some studies reported higher prevalence among boys [33], while others found girls at greater risk [37]. Understanding the cultural and behavioural reasons for these differences is important. Caregiver practices, feeding routines, and attitudes toward hygiene may vary by sex due to deeply ingrained social norms [38–40]. Ethnographic and qualitative research can help explain these patterns and inform interventions that are sensitive to cultural context.

Second, the study findings also suggest the possible impact of systemic inequities limiting access to dental care. The current study indicates that between 50% and 95% of children have never visited a dentist. The severe shortages of dental personnel in the region may have contributed to this. The region has only 0.03 dentists per 10,000 population, no dental assistants or therapists, and 0.17 dental prosthetic technicians per 10,000 population [9]. These gaps, together with the lack of universal oral health coverage and the absence of robust policies and programmes [37, 41] may contribute to the high prevalence of ECC. In addition, the Central Africa Republic and the Democratic Republic of Congo face humanitarian crises [42–44]; the ECC prevalence may reflect disrupted health systems. Past studies had shown that wars and humanitarian crisis increases the risk for ECC [45] There is also the possible contribution of culture to poor dental service utilization [46]as the availability of services does not always translate to their uptake [47]The findings on the low prevalence of ECC among Baka Pygmies are a pointer to the possibility of protective cultural/dietary practices. Further studies are needed to understand the context-specific drivers of the high prevalence of ECC in the region.

Third, the high prevalence of ECC in the region has significant implications. Children in the region are highly vulnerable to a preventable oral health condition that undermines nutrition, growth, learning, and overall wellbeing [2–7]. The lack of integration of oral health into primary healthcare and universal coverage frameworks in countries in the region may further delay early detection and treatment [9]. As a result, ECC becomes not only a clinical burden but also a marker of broader systemic health inequities that demand urgent attention.

Interestingly, one study reported that children whose mothers had experienced dental caries were less likely to develop ECC [33]. This suggests that prior personal experience of dental disease could potentially lead to protective caregiving behaviour. However, this observation requires cautious interpretation as this was based on a single study.Further investigation is needed to confirm causality.

Fourth, the study findings suggest that addressing ECC in Central Africa would require action at multiple levels. Interventions should target individual behaviour, caregiver knowledge, school environments, and health systems. Integrating ECC prevention into existing maternal and child health programs [48], establishing school-based oral health promotion initiatives [18], and training mid-level oral health providers [9]. These are all important strategies that can contribute to reducing the prevalence of ECC in the region. Community-based education and partnerships with traditional caregivers could further strengthen prevention efforts and improve uptake of preventive oral health services. For a region where the oral healthcare system appears fragile, a strong focus on preventive oral health care may be critical; otherwise, the prevalence of ECC may continue to increase into the future.

The wide confidence interval of the pooled prevalence, the confirmation of an outlier-driven instability, and the variability in the tools used to measure ECC suggest that the study result should be taken with caution. The study findings can, at best, drive the development of hypotheses rather than inform policies. Future research should prioritize generating information about the epidemiological profile of ECC in the countries in the region with no data and designing studies with less risk of potential bias. These studies could adopt longitudinal and community-based designs to better assess the incidence and causal pathways of ECC. Qualitative studies exploring caregivers’ beliefs and practices will also be critical to inform culturally appropriate interventions. Furthermore, health policy research should evaluate the feasibility and cost-effectiveness of regional ECC prevention strategies, such as fluoride varnish application and sugar reduction campaigns.

## Conclusion

This study reveals a high prevalence of ECC in selected countries in Central Africa, affecting more than half of children under six years of age. Socioeconomic inequalities and cultural factors may facilitate the risks of ECC in the region. The paucity of robust data may impede effective policy and programmatic responses. Addressing the ECC burden in the region requires actions that can promote the integration of ECC prevention into maternal and child health services, school-based oral health programs, and regionally tailored policies to expand access to affordable dental care. Strengthening research in currently underrepresented countries is also crucial to fully characterize and combat ECC across the subregion.

## Declarations

### Patients and Public Involvement

Patients and members of the public were not involved in the design, conduct, reporting, or dissemination plans of this systematic review and meta-analysis because the study was based exclusively on analysis of previously published literature.

### Ethics approval and consent to participate

Not applicable

### Consent for publication

Not applicable

### Availability of data and materials

All data used in the study are available to the public

### Authors’ contributions

OO: Conducted the search, screened title and abstract, extracted data, risk of bias assessment, conducted the meta-analysis, and drafted the manuscript and reviewed the manuscript for intellectual inputs.

CO: Conducted the search, title and abstract screening, full text screening, and data extraction, risk of bias assessment

FA: Generated search terms, protocol preparation, and reviewed the manuscript for intellectual inputs.

AE: Conceptualization of the work, protocol review, and reviewed the manuscript for intellectual inputs

GUE: Conceptualization, protocol review and reviewed the manuscript for intellectual inputs

QOL: Conceptualization, protocol review and reviewed the manuscript for intellectual inputs

RO: Search term generation and review, meta-analysis, and reviewed the manuscript for intellectual inputs

OCE: Conceptualization, Protocol review, and reviewed of the manuscript for intellectual inputs

AMA: Technical support, search for grey literature, protocol review, manuscript review and reviewed of the manuscript for intellectual inputs

MOF: Conceptualization, protocol review, manuscript writing and review, data extraction, technical support, project management.

## Data Availability

All articles included in the review are available on line and referenced in the manuscript

## Acknowledgments

We would like to thank the Oral Health Initiative for its contributions to the success of this systematic review. We would also like to thank Drs. Wale Adejobi, Samuel Ajigbotosho, and Ayobami Bakare for their contributions to the success of this review.

## List of abbreviations

ECC: Early Childhood Caries
PEO: Population, Exposure, and Outcomes
PROSPERO: Prospective Register of Systematic Reviews
PRISMA: Preferred Reporting Items for Systematic Reviews and Meta-analyses

## PUBMED

**Supplemental File 1:**
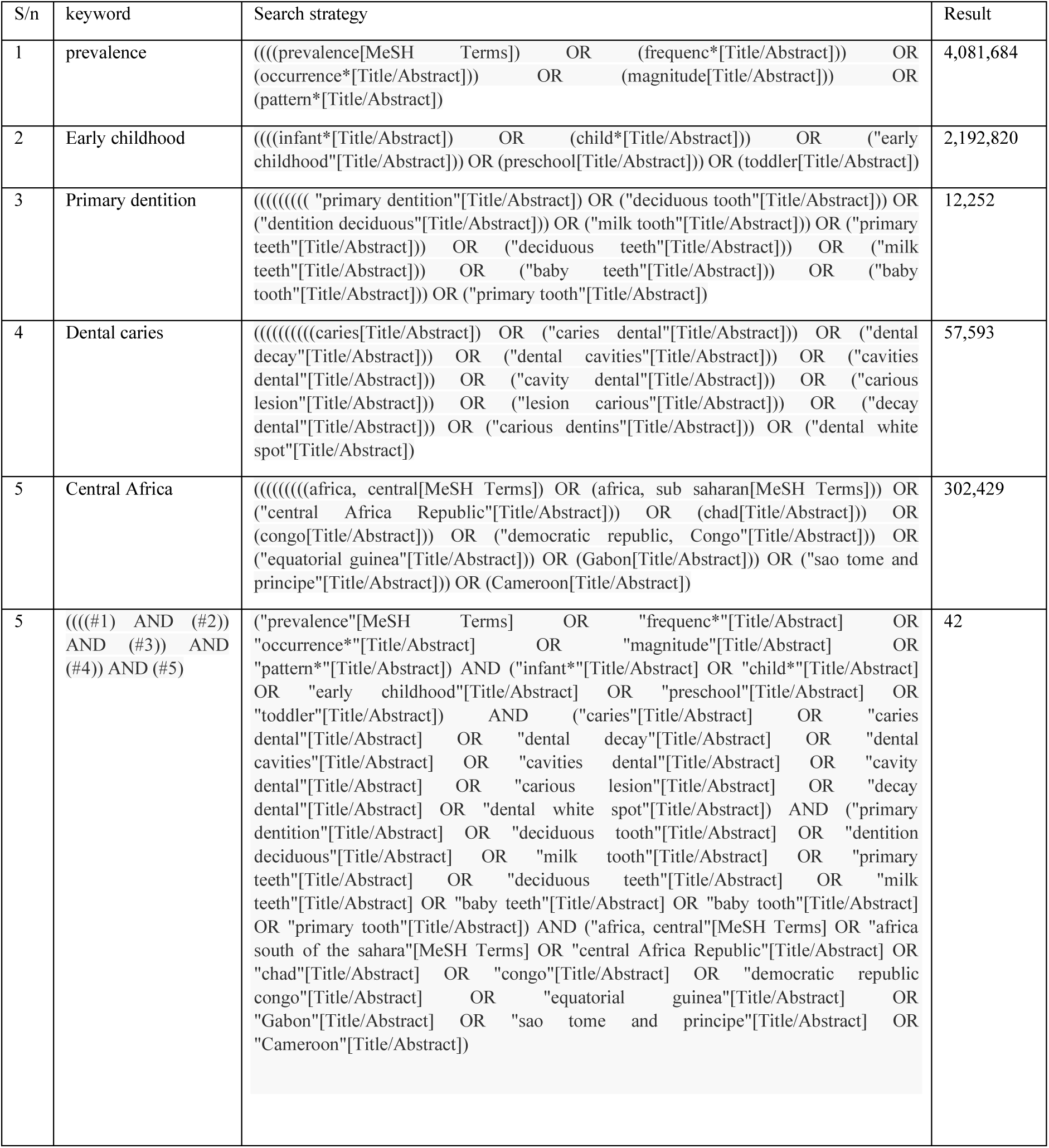
Search Strategy for Prevalence and associated Risk Factor of Early Childhood Caries in Central Africa; A Systematic Review and Meta Analysis PUBMED.

## Proquest

summary(Prevalence OR frequenc* OR occurrence* OR magnitude OR pattern* OR burden) AND summary(infant* OR child* OR “Early Childhood” OR preschool OR toddler) AND summary(caries OR “caries dental” OR “dental cavity” OR “dental decay” OR “dental cavities” OR “cavities dental” OR “cavity dental”OR “carious lesion” OR “lesion carious” OR “decay dental” OR “carious dentins”OR “dental white spot) AND summary(Cameroon OR “central Africa Republic” OR chad OR Congo OR “democratic republic Congo” OR “equatorial guinea” OR Gabon OR “sao tome and principe)” =3

## SCOPUS

TITLE-ABS-KEY (Prevalence OR frequenc* OR occurrence* OR magnitude OR pattern* OR burden) AND TITLE-ABS-KEY (infant* OR child* OR “Early Childhood” OR preschool OR toddler) AND TITLE-ABS-KEY (caries OR “caries dental” OR “dental cavity” OR “dental decay” OR “dental cavities” OR “cavities dental” OR “cavity dental” OR “carious lesion” OR “lesion carious” OR “decay dental” OR “carious dentins” OR “dental white spot”) AND TITLE-ABS-KEY (“primary dentition” OR “deciduous tooth” OR “dentition deciduous” OR “milk tooth” OR “primary teeth” OR “deciduous teeth” OR “milk Teeth” OR “baby teeth” OR “baby tooth” OR “primary tooth”) AND TITLE-ABS-KEY (cameroon OR Chad OR Congo OR “Democratic Republic, Congo” OR “equatorial Guinea “ OR “gabon” OR “sao tome and Principe” OR “central Africa Republic”)) =1

## CINAHL

AB (Prevalence OR frequenc* OR occurrence* OR magnitude OR pattern* OR burden) AND AB ((infant* OR child* OR “Early Childhood” OR preschool OR toddler) AND AB (caries OR “caries dental” OR “dental cavity” OR “dental decay” OR “dental cavities” OR “cavities dental” OR “cavity dental”OR “carious lesion” OR “lesion carious” OR “decay dental” OR “carious dentins”OR “dental white spot) AND AB (Cameroon OR “central Africa Republic” OR chad OR Congo OR “democratic republic Congo” OR “equatorial guinea” OR Gabon OR “sao tome and principe) =16

## AJOL

(Prevalence OR frequency* OR occurrence* OR magnitude OR pattern* OR burden) AND (infant* OR child* OR “Early Childhood” OR preschool OR toddlers) AND (“primary dentition*” OR “deciduous tooth” OR “dentition deciduous” OR “mild tooth” OR “primary teeth” OR “deciduous teeth” OR “mild Teeth” OR “baby teeth” OR “baby tooth” OR “primary tooth”) AND (caries OR “caries dental” OR “dental cavity” OR “dental decay” OR “dental cavities” OR “cavities dental” OR “cavity dental”OR “vicarious lesion” OR “lesion vicarious” OR “decay dental” OR “vicarious dentins” OR “dental which spot”) AND (Cameroon OR “central Africa Republic” OR chan OR Congo OR “democratic republic Congo” OR “equatorial guinea” OR Gabon OR “sao tome AND principe”) = 9

## WEB OF SCIENCE

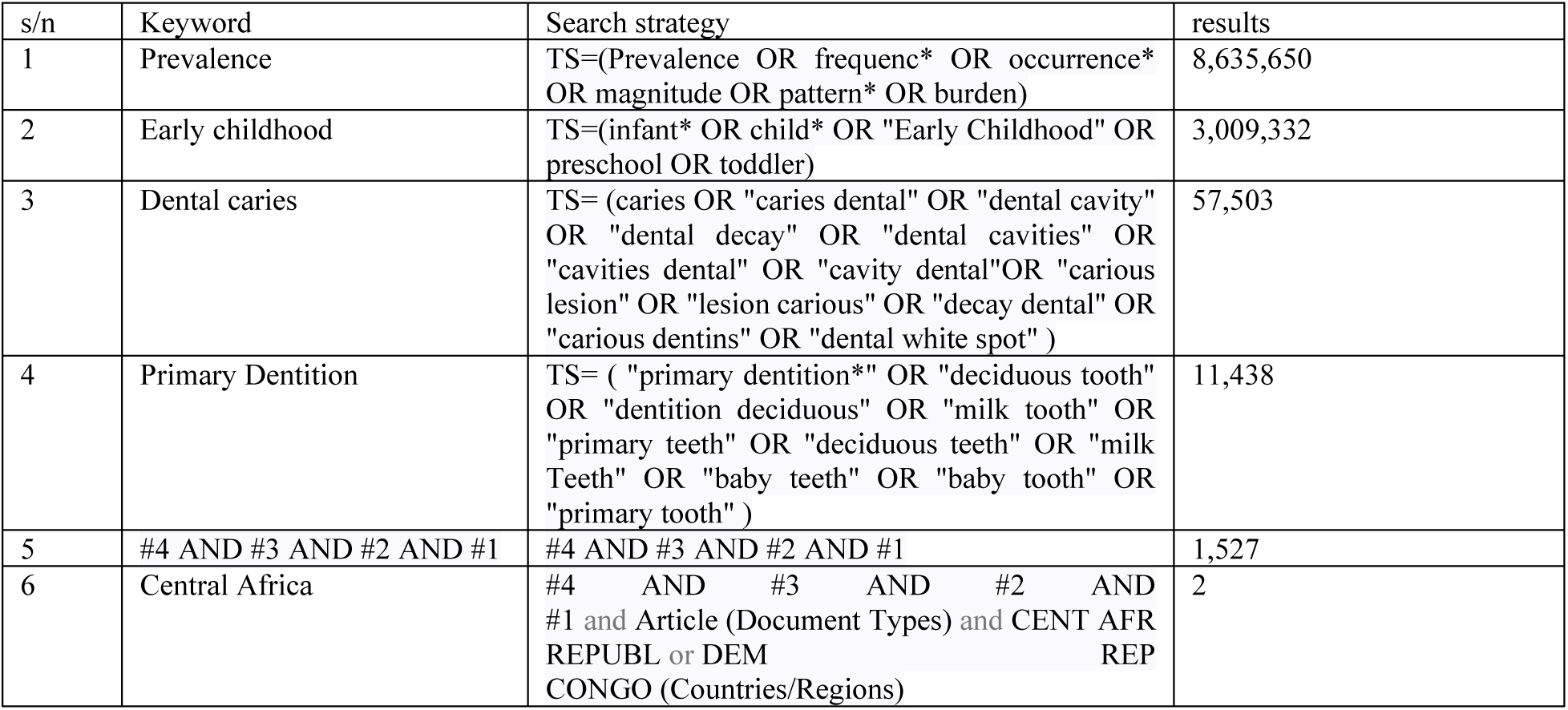

**Supplemental File 2:**
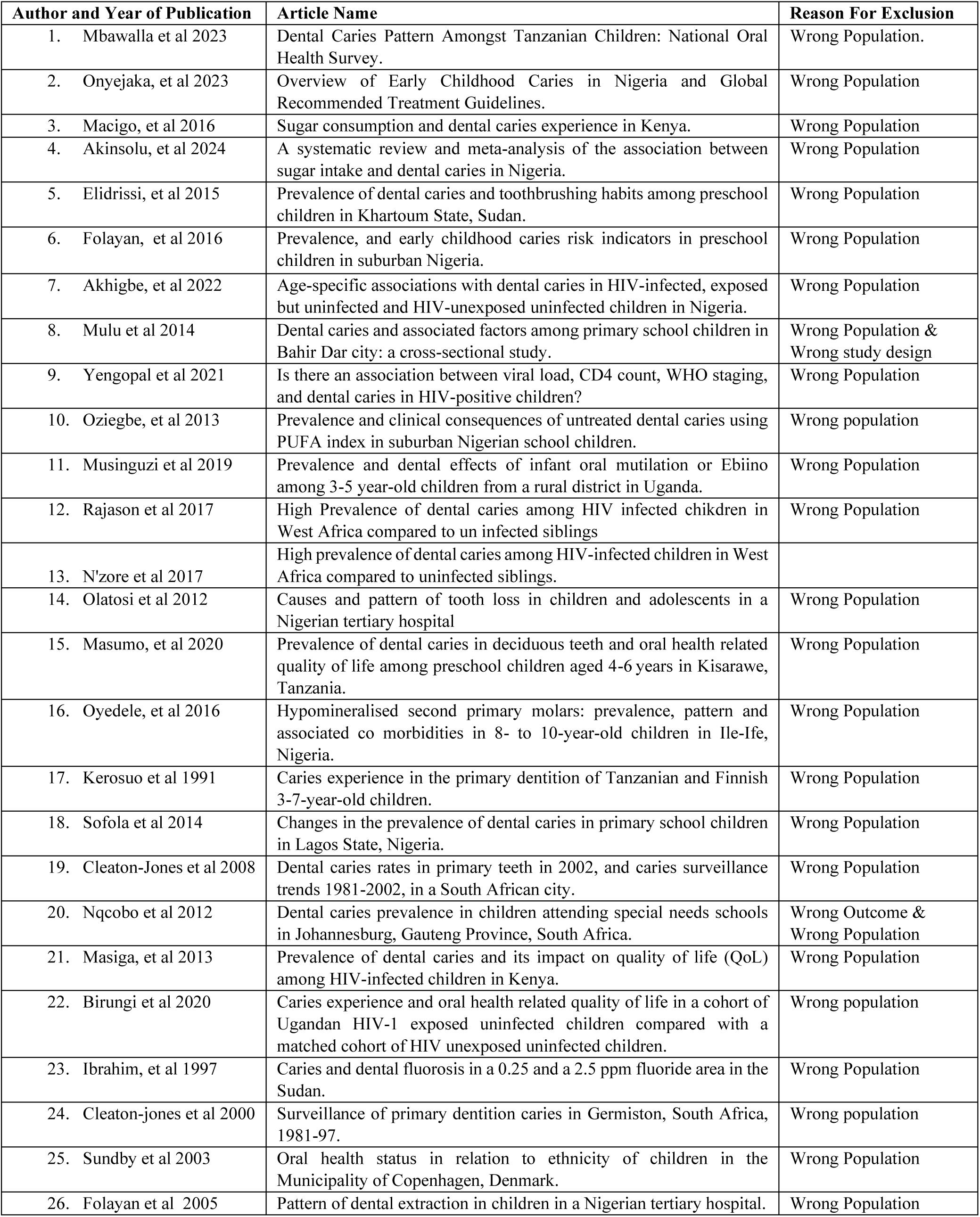
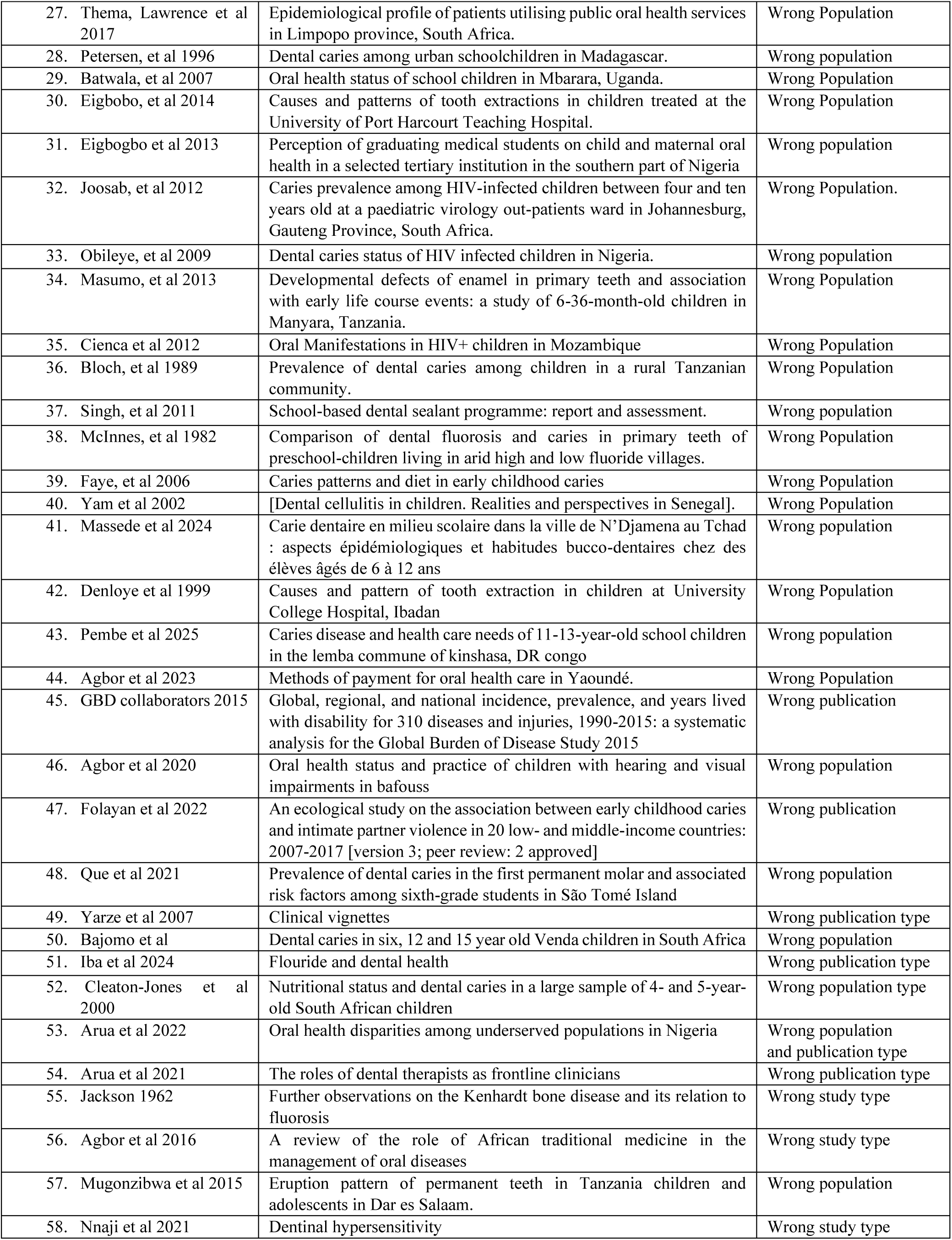
Excluded Studies and Reasons for Exclusion.

**Supplementary File 3.** Inter-rater Reliability Assessment.

| Assessors | Reference | Were the criteria for inclusion in the sample clearly defined? | Were the study subjects and the setting described in detail? | Was the exposure measured in a valid and reliable way? | Were objective, standard criteria used for measurement of the condition? | Were confounding factors identified? | Were strategies to deal with confounding factors stated? | Were the outcomes measured in a valid and reliable way | Was appropriate statistical analysis used? |
| --- | --- | --- | --- | --- | --- | --- | --- | --- | --- |
| Reviewer A | Aimond et al [30] | 2 | 2 | 2 | 2 | 0 | 1 | 2 | 0 |
| Reviewer B | Aimond et al [30] | 2 | 2 | 2 | 1 | 1 | 1 | 2 | 2 |
| Reviewer A | Lamloum et al [34] | 2 | 2 | 2 | 2 | 2 | 1 | 2 | 2 |
| Reviewer B | Lamloum et al [34] | 2 | 2 | 2 | 2 | 2 | 1 | 2 | 2 |
| Reviewer A | Songo et al [33] | 2 | 2 | 2 | 2 | 2 | 2 | 2 | 2 |
| Reviewer B | Songo et al [33] | 2 | 2 | 1 | 1 | 2 | 1 | 2 | 2 |
| Reviewer A | Kuenang [31] | 2 | 1 | 2 | 2 | 0 | 0 | 2 | 0 |
| Reviewer B | Kuenang [31] | 2 | 2 | 1 | 2 | 2 | 2 | 2 | 2 |
| Reviewer A | Poni et al [32] | 2 | 2 | 2 | 2 | 0 | 1 | 2 | 2 |
| Reviewer B | Poni et al [32] | 2 | 2 | 2 | 2 | 2 | 1 | 2 | 2 |
| Percentage Agreement | Overall (70%) | 100% | 80% | 60% | 60% | 40% | 60% | 100% | 60% |

